# Microbial Cell-Free DNA Sequencing for Diagnosing Lung Infections in Immunocompromised Patients

**DOI:** 10.1101/2025.11.24.25340574

**Authors:** Kevin Brick, Kristin H. Jarman, Frederick Nolte, Igor D. Vilfan, Marla Lay Vaughn, Aga Zielinska, Victoria Portnoy, Bradley A. Perkins, Sivan Bercovici, Timothy A. Blauwkamp, Jason D. Goldman, Joshua A. Hill, Stephen P. Bergin

**Affiliations:** Karius, Inc. 975 Island Drive, Redwood City, CA 94065; Providence Swedish Medical Center, 1124 Columbia St. #600, Seattle, WA, 98144; Division of Allergy and Infectious Diseases, University of Washington, 1959 NE Pacific Street, Seattle, WA, 98195; Vaccine and Infectious Disease Division, Fred Hutchinson Cancer Center, 1100 Fairview Ave. N., Mail Stop E4-100, Seattle, WA 98109, USA; Duke Clinical Research Institute, Duke University School of Medicine, DUMC 2629, 203 Research Dr MSRB1 Ste. 201B, Durham, NC 27710

## Abstract

**Rationale:** Rapid and comprehensive detection of respiratory pathogens is essential for immunocompromised patients with suspected pneumonia. Conventional diagnostic methods are limited in scope, often slow, and lack sensitivity, leading to reduced diagnostic yield.

**Objectives:** To analytically and clinically validate a microbial cell-free DNA (mcfDNA) sequencing assay for bronchoalveolar lavage fluid (BALF) that enables broad and sensitive detection of pathogens causing pneumonia in immunocompromised patients.

**Methods:** The BALF-mcfDNA assay detects more than 500 bacteria, DNA viruses, fungi, and parasites. Analytical validation used 324 contrived samples containing DNA from 11 representative organisms and 1,294 in-silico samples. Clinical validation compared BALF-mcfDNA sequencing with usual care (UC) testing in 118 immunocompromised adults with suspected pneumonia. An independent expert panel adjudicated the etiologic diagnoses.

**Measurements and Main Results:** BALF-mcfDNA testing demonstrated high inclusivity (97%), exclusivity (96%), and precision (coefficient of variation <15%), with linear quantification across four orders of magnitude. Limits of detection were consistent across taxa, GC content, and genome size. In clinical evaluation, BALF-mcfDNA sequencing identified probable pneumonia pathogens in 34% of participants versus 24% for UC testing, increasing diagnostic yield by 43% (57% when combined with UC). In 24% of concordant cases, mcfDNA sequencing provided additional clinically relevant information by identifying new or species-level pathogens.

**Conclusions:** BALF-mcfDNA sequencing enables sensitive, comprehensive, and rapid pathogen detection from BALF specimens, outperforming conventional methods and improving etiologic diagnosis of pneumonia in immunocompromised hosts.

## Introduction

Lower respiratory tract infections cause significant morbidity and mortality worldwide, especially among the millions of patients immunocompromised due to cancer treatments, biological therapies, organ transplants, or immune disorders(1). Most pneumonia cases in these patients lack pathogen identification(2), which often necessitates empirical treatment that may lead to antimicrobial resistance and sub-optimal outcomes. Advanced respiratory diagnostics with greater sensitivity, less bias, and faster pathogen detection are critically needed(3).

Bronchoalveolar lavage (BAL) is used to obtain lower respiratory samples for microbiological analysis when infection is suspected, but the diagnostic yield from BAL fluid (BALF) with current diagnostics is often low, especially after antimicrobial exposure, which suppresses pathogen growth *in-vitro*. In immunocompromised patients, who may harbor diverse and hard-to-detect pathogens, an extensive battery of diagnostic testing is often needed to identify the causative agent, and still produces sub-optimal diagnostic yield(1).

To address diagnostic gaps in conventional methods and consolidate diagnostic tests, various culture-independent tests, such as syndromic PCR panels(4, 5) and targeted sequencing(6), have been developed, but these remain limited to certain taxa. Pathogen-agnostic metagenomic sequencing(7–9) potentially offers comprehensive, unbiased, high-throughput pathogen detection. Microbial cell free DNA (mcfDNA) fragments, released by immune attack, microbial autolysis and possibly treatment, allow detection of microbial DNA without cell disruption. Thus, mcfDNA sequencing can minimize disruption to commensal taxa, streamline sample processing and shorten analysis time compared to whole-cell-DNA mNGS methods(10). In a prospective multicenter study of 173 immunocompromised adults undergoing bronchoscopy for pneumonia (PICKUP study), plasma-mcfDNA sequencing increased the relative diagnostic yield by 40.2% when added to usual care testing(2).

We hypothesized that metagenomics analysis from mcfDNA in BALF would also increase the diagnostic yield over usual care BALF-based diagnostics, however, applying clinical metagenomics to non-sterile sites like the lungs poses challenges, including distinguishing true pathogens from commensal flora and interpreting results that vary due to differences in patient populations and sample collection(11). Here, we report the validation of a novel metagenomic sequencing test for identifying bacterial, DNA viral, fungal, and parasitic pathogens from mcfDNA in BALF. The analytical validation addresses these unique challenges of metagenomic testing with a multi-faceted strategy combining real-world clinical samples, strategically designed contrived specimens, and *in-silico* experiments. Clinical validation addressed clinical performance in a highly phenotyped clinical trial cohort, using residual BALF from the PICKUP study(2). By leveraging the power of metagenomic sequencing, this test has potential to substantially increase diagnostic yield and improve outcomes in patients with suspected lower respiratory tract infections.

## Methods

### Bronchioalveolar lavage fluid microbial cell free DNA sequencing (Karius Focus^TM^|BAL)

Metagenomic testing of mcfDNA in BALF (BALF-mcfDNA sequencing / Karius Focus|BAL) was performed at Karius Inc., a CLIA-certified, CAP-accredited, and New York State Department of Health-approved laboratory. The Clinical Reportable Range (CRR) of the test contains >500 microbial taxa with potential to cause lung infections, classified as *Obligate and Opportunistic Pathogens* (may cause lung infection), *Microbes with Pathogenic Potential and DNA Viruses* (may cause lung infection or represent commensals), *Upper Respiratory Tract Flora* (more commonly commensals than the cause of lung infection). To detect microbes, metagenomic sequencing was performed from BALF using the Helion in-matrix method(10) and a workflow adapted from plasma mcfDNA sequencing(12) (Fig. 1A). Sequencing data were processed similarly to data from plasma mcfDNA sequencing(12). Extensive methodological details are provided in the online supplementary materials.

**Figure 1.**
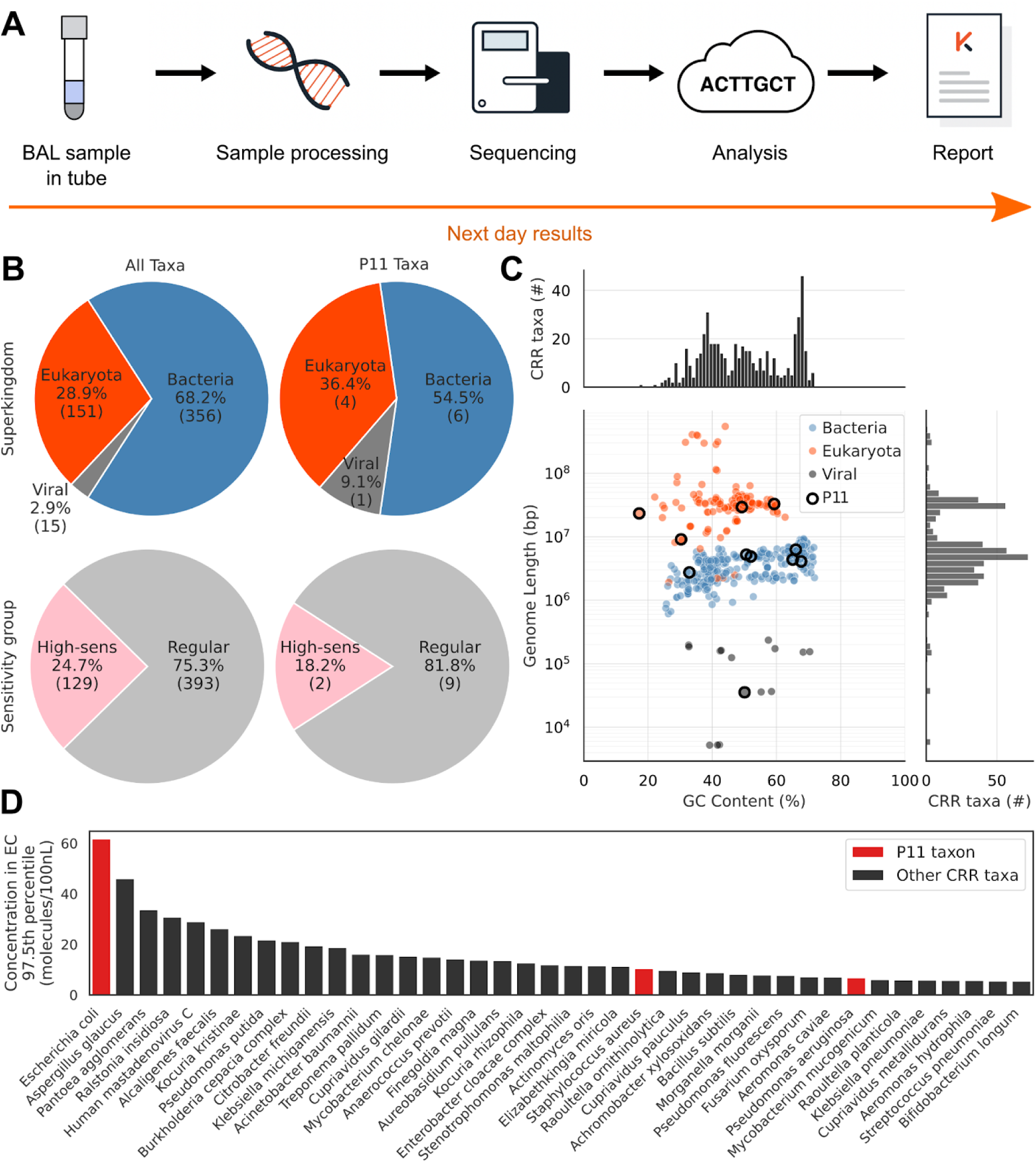
Contrived pathogen mix is representative of the BALF-mcfDNA test CRR. A reference panel of 11 microorganisms was designed to reflect the diversity of the 522 microorganisms tested by the assay. **A.** A schematic overview of the BALF-mcfDNA sequencing process. **B.** Relative number of microorganisms comprising the CRR or contrived taxa by superkingdom (top), and by sensitivity group (bottom). **C.** The GC-content and genome length of all microorganisms comprising the CRR are shown, colour coded according to superkingdom.

### Analytical validation

Genomic DNA from 11 representative CRR microorganisms spanning genome size and GC content was fragmented (to reflect mcfDNA fragment lengths in BALF) and pooled (P11 mixture). P11 were added to defined background matrices prior to sequencing. Details are provided in the online supplementary materials. Following CLSI EP17-A2(13), the LoD was calculated using probit analysis for each P11 taxon spiked into BALF at 13 concentrations. The reportable range was defined by the lower and upper limits of quantitation (LLoQ/ULoQ)(12).

Sequencing reads were simulated *in-silico* from microbial assemblies to evaluate assay inclusivity, exclusivity, cross-reactivity, and informatics stability. For inclusivity and exclusivity, simulated reads from blinded assemblies tested the impact of sequence divergence and off-target taxa. Informatics stability assessed pipeline performance for all CRR taxa.

Cross-reactivity assessed the impact of closely related taxa on microbial calls and concentration estimates. Quantitative accuracy compared automated electrophoresis (TapeStation, Agilent, Santa Clara, CA) DNA concentration estimates to estimates from BALF-mcfDNA sequencing, using criteria that satisfy CLSI EP09-A3(14). Within-run and between-run precision were assessed using P11 spiked into human BALF reference matrix at concentrations above LoD, and in accordance with CLSI EP05-A3 guidelines(14). Extensive methodological details for validation experiments are provided in the online supplementary materials.

### Clinical validation

Clinical validity was assessed using retrospective data from a prospective, multicenter study at ten U.S. tertiary care centers (PICKUP, NCT04047719)(2, 15). Hospitalized patients undergoing bronchoscopy for suspected pneumonia were eligible if they were immunocompromised due to hematologic malignancy, hematopoietic cell transplantation, or immunosuppressive therapy for graft-versus-host disease. Of 249 enrolled participants(2), 118 met inclusion criteria (Fig. S5).

Participants received standard diagnostic testing (“usual care,” UC) that included BALF bacterial, fungal, and mycobacterial cultures; *Pneumocystis jirovecii* testing; blood cultures; serum galactomannan; and respiratory viral panels. BALF-mcfDNA sequencing results were compared with UC to determine diagnostic sensitivity, specificity, and additive yield.

An expert adjudication panel of nine clinicians reviewed UC and BALF-mcfDNA sequencing results, along with relevant clinical data, to identify the most likely causes of pneumonia. Each case was independently reviewed by two adjudicators, with a third resolving disagreements. Detailed study design, inclusion/exclusion criteria, and adjudication procedures are provided in the online supplementary materials.

## Results

### Validation strategy

We validated the BALF-mcfDNA sequencing test using a Clinical & Laboratory Standards Institute (CLSI) aligned strategy that addressed the unique challenges of clinical metagenomics. Key to this validation was a contrived sample panel of 11 taxa (P11) representing the diversity, superkingdoms, genomic features, and environmental contaminant profiles of the reportable taxa (Fig. 1B-D). To complement the P11 strategy, 1,294 *in-silico* samples covering genetic diversity and co-infection scenarios and 42 clinical samples to assess precision were analyzed. Clinical validation examined 118 clinical study samples from immunocompromised patients with suspected pneumonia to compare to the best usual care standard.

### Analytical performance characterization

The limit of detection (LoD) was determined through experiments with P11 samples that accounted for key technical variables affecting sensitivity, including sequencing depth, human DNA content and microbe-specific differences. The P11 taxa, comprising 55% bacteria, 36% eukaryota, and 9% viruses were chosen to reflect the major microbial superkingdoms on the CRR (68%, 29%, and 3%, respectively; Fig. 1B). Likewise, their GC content and genome lengths were comparable to those of CRR taxa (Fig. 1C). Reference BALF samples with P11 DNA spiked at varying concentrations were used to calculate LoD using probit fits (Fig. 2A; Table S2). By evaluating LoD across typical sequencing sample depths, we observed that the LoD was modestly higher at low depth (Fig. 2B); LoD ranged from 304 to 45 molecules/100nL for low-EC taxa, and from 1,138 to 248 molecules/100nL for regular-EC taxa. *E. coli* showed slightly elevated LoD at shallow depths due to environmental contamination (Fig. 1D). Global LoDs were 153 and 595 molecules/100nL for low EC-risk and regular EC-risk taxa, respectively. LoD estimates were similar for microbes within each EC-risk category (Fig. 2B; Table S2) with no correlation to genomic GC content or genome length (Fig. S1). Human cfDNA concentration had minimal impact; samples at the 90th percentile exhibited only 17-23% LoD increase (Fig. S2).

**Figure 2.**
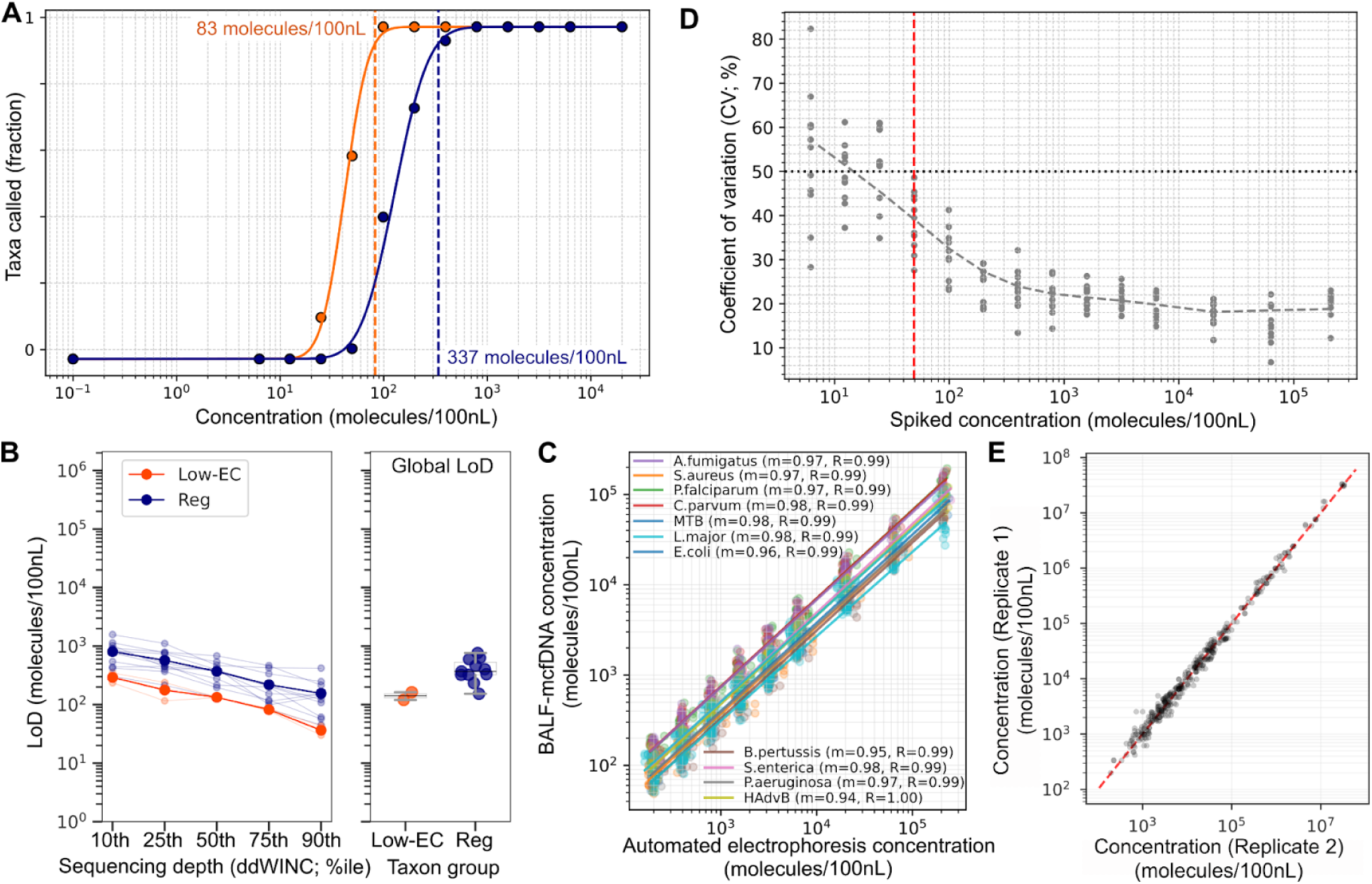
Analytical validation of the BALF-mcfDNA test. **A.** An example of the probit fit used to calculate LoD for high-sensitivity (orange) and regular-sensitivity (navy) taxa. Data are from the LoD calculated at the 75th percentile of sequencing depth. **B.** The LoD was determined for each of the 11 taxa in the contrived pathogen mix across the range of sequencing depths observed in pre-validation clinical samples. (left panel) Individual taxa are shown as faded lines colored by Low-EC risk and regular EC risk taxa. Taxa were pooled and the LoD calculated for each group (solid lines). (right panel) The global LoD was also calculated for each taxon and taxon group. **C.** BALF-mcfDNA concentration estimates linearly scale with concentration estimates of DNA concentration from automated electrophoresis. Each point represents one measurement for a P11 taxon. The slopes (m) and correlation coefficients (R) are shown for each taxon in the legend. **D.** Pipeline concentration estimates are stable below the LoD. The lower limit of quantitation was set at 50 molecules/100nL (red line), the closest integer value above the concentration at which the CV does not exceed 50% for any taxa. Each data point represents the coefficient of variation of concentrations across all replicates for a single microbe at the indicated spiked concentration. **E.** High precision concentration estimates for reportable taxa in clinical samples. All pairwise comparisons of samples run in triplicate are shown. Each point represents a single taxon. The microorganisms used for contrived reference materials are shown as black circles. Histograms above and to the right of the scatter plot show the total number of CRR organisms at each GC-content and genome length. **D.** The 97.5th percentile abundance observed in 136 negative control samples from validation environmental controls is shown for the 40 CRR microorganisms found as top environmental contaminants (only taxa with >5 detections are shown). The red bars indicate the microorganisms that are part of the contrived reference materials.

Strong linearity and quantitative accuracy was observed for concentration estimates across four orders of magnitude for all P11 taxa (Fig. 2C; Fig. S3). Notably, BALF-mcfDNA concentrations were ∼50% lower than DNA concentrations by electrophoresis, indicating a systematic shift between methods. BALF-mcfDNA sequencing gave precise relative concentrations, even below the global LoD (Fig. 2D), and the LLoQ was established at 50 molecules/100nL (Fig. 2D). The ULoQ was 210,000 molecules/100nL (the highest concentration tested), based on linearity with electrophoresis estimates. Thus, the quantitative reportable range is 50-210,000 molecules/100nL (Fig. 2C).

Quantitative precision was determined from the CV of mcfDNA concentrations under CLSI-aligned conditions (see methods). Using contrived samples, the within-run and between-run variability were 7.4% and 13.8%, respectively (Table S3). The between-run variability for pairwise observations in clinical samples was 14.8% (Fig. 2E), with 93.8% pairwise call concordance. As expected, missing calls were subject to analytical filtering or close to LoD with p-values just short of significance.

Analytical specificity is paramount for clinical metagenomic assays, as contaminating DNA from environmental, reagent, or cross-sample sources can yield misleading results. To assess the analytical specificity of BALF-mcfDNA sequencing under clinically relevant conditions, we examined 72 AC samples and found no unexpected taxa. Thus, estimated analytical specificity is >95.9% (0/72; 100% [95% CI: 95.9-100%]).

*In-silico* simulations allowed assessment of test performance beyond practical lab limits. We first verified that all CRR taxa could be reported from simulated sequencing reads (522/522; PPA = 100% [95% CI:99.3-100%].). 99.2% of concentration estimates were within 20% of expected values (Fig. 3A). In simulations to test the impact of genetic variation (inclusivity; see methods), the correct taxon was reported in 103/106 cases (PPA = 97.2% [95% CI: 92.0-99.4%]; Fig. 3A, Table S4), and only the expected taxon was reported in 102/105 positives (PPV = 98.1% [95% CI:93.3-99.8%]). Four simulations reported one incorrect on-CRR taxon from the same family.

**Figure 3.**
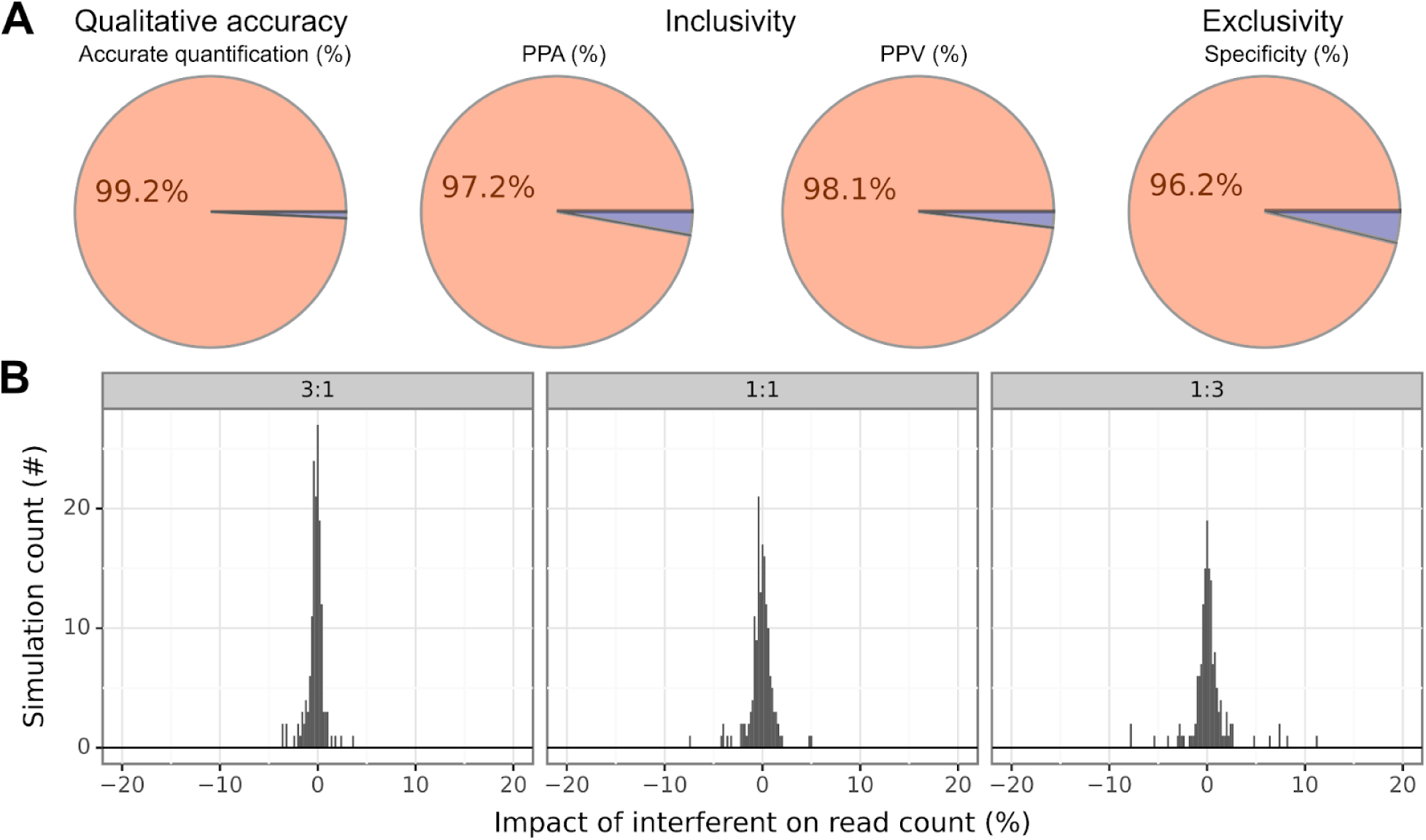
*In-silico* experiments demonstrate the robustness of the BALF-mcfDNA test to natural genetic variation. **A.** *In-silico* simulations demonstrate that the test can accurately detect and quantify the concentration of all taxa on the CRR (Informatics stability; Accurate quantification). *In-silico* simulations also show that the presence of very closely related species does not have a large impact on BALF-mcfDNA test results (Inclusivity & Exclusivity). **B.** *In-silico* interference experiments demonstrate that quantification is minimally impacted by the presence of closely related taxa in a sample. 96.2% of comparisons show less than a 20% impact on the target taxon read count when a related interfering taxon is present (experiments with values outside this range are not shown here). Data are shown for each target taxon : interfering taxon ratio (3:1, 1:1, 1:3). Note that the percentage of simulations with an impact close to zero is greatest when the target taxon is in excess (3:1).

For similar experiments using non-CRR taxa (exclusivity; see methods), no taxa were reported in 102/106 simulations (specificity = 96.2% [95% CI: 90.6-99.0%]; Fig. 3A), with four false positives in closely related species of the same genus (Table S5). We also evaluated cross-reactivity by spiking two related taxa at different ratios (3:1, 1:1, 1:3). Despite the presence of related taxa, the mean EDR was 96.6% of expected (95% CI: 96.5-96.7%; N=480), with 96.2% of pairs showing <20% deviation (Fig. 3B). Larger deviations primarily involved three highly similar taxon pairs (Table S6).

### Clinical Validation

We employed external adjudication to compare the clinical performance of BALF-mcfDNA sequencing with UC-testing in 118 immunocompromised patients with suspected pneumonia(2, 15) (Table S7; adjudication details in methods). Tables S7 & S8 provide summaries of the cohort demographics, including testing results and adjudications.

#### Clinical sensitivity

BALF-mcfDNA sequencing identified an adjudicated cause of pneumonia in 34% (40/118) of cases, significantly more often than individual UC tests (0-8%; P<10^-5^ Fischer’s exact test) or composite-UC testing (24% (28/118); P=0.01 McNemar Test) (Fig. 4A-B). Aspergillus PCR showed no significant difference, but power was limited by small sample size (N=12). Among samples with an adjudicated cause of infection, BALF-mcfDNA sequencing missed fewer detections (4 patients) than composite-UC (16 patients), and provided additional (6 patients) or more specific (6 patients) pathogen information than UC in 12 patients, including species-level identification for *Aspergillus, Nocardia*, and *Mucorales* (see Tables S9-S12). Thus, the diagnostic yield (the proportion of participants with an adjudicated etiology in which each test identified the relevant microbe) of BALF-mcfDNA sequencing (40/44; 91% [95% CI: 82-100%]) was 43% higher than that of composite-UC testing (28/44; 64% [95% CI: 53-75%]) (p=0.01; McNemar Test). Of 82 taxa adjudicated to cause pneumonia in 44 patients, 57% (47/82) were detected only by BALF-mcfDNA, 12% (10/82) only by composite-UC, and 30% (25/82) by both (Fig. 4C; Table S8). Overall, BALF-mcfDNA sequencing significantly improved the identification of the cause of lung infections (P=0.01 McNemar Test), providing exclusive or additional clinically relevant findings in 24% of patients. Adding BALF-mcfDNA sequencing to composite-UC resulted in a 57% increase in diagnostic yield compared to composite-UC alone.

**Figure 4.**
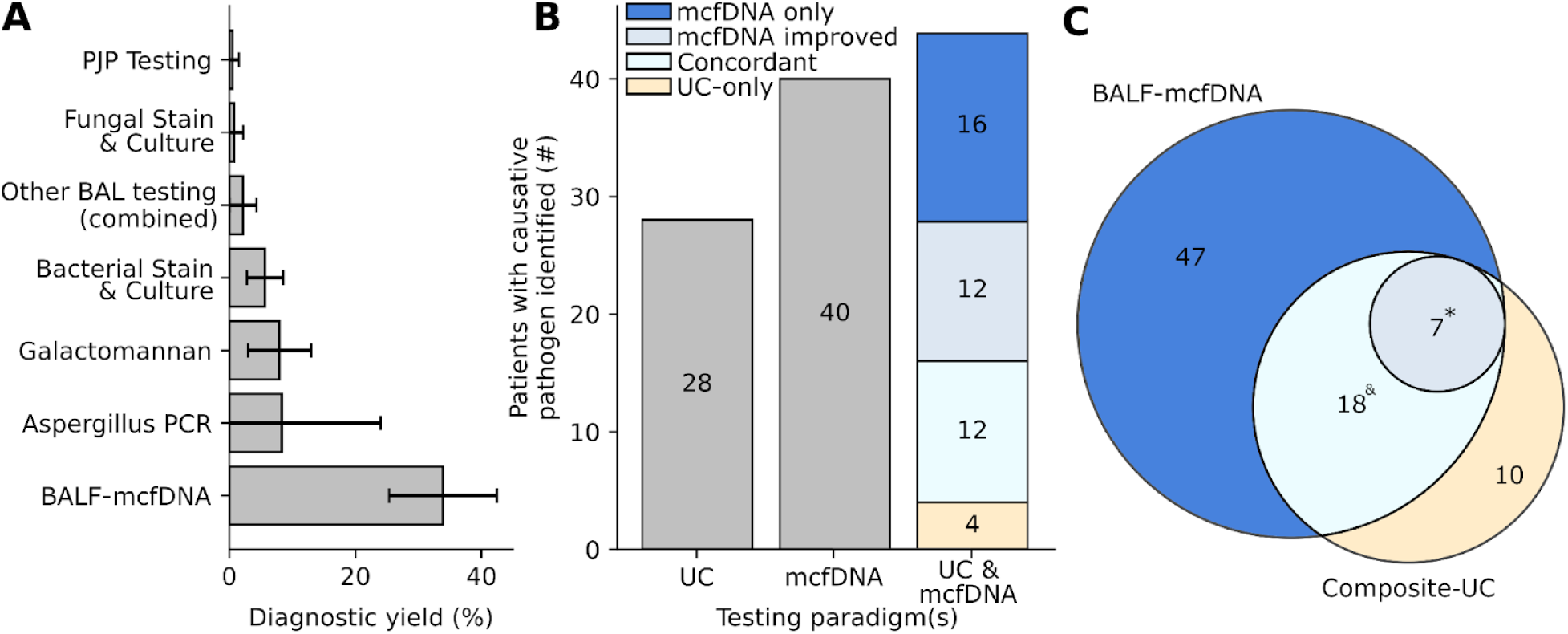
BALF-mcfDNA sequencing improves clinical diagnoses compared to composite-UC testing. **A**. The diagnostic yield was measured as the percent of tests identifying an adjudicated cause of pneumonia across all cases. No other usual care tests identified adjudicated causes of pneumonia in this patient cohort. Error bars indicate 95% confidence intervals. **B.** An adjudicated cause of pneumonia was identified in 12 more patients using BALF-mcfDNA sequencing (mcfDNA) than using composite-UC (UC) testing. BALF-mcfDNA sequencing also yielded additional diagnostic information in 12/24 patients in whom both testing paradigms identified a concordant cause of pneumonia (mcfDNA improved). **C.** Per-microbe comparison of adjudicated causes of pneumonia. BALF-mcfDNA sequencing identified 47 microbes causing pneumonia that were missed by composite-UC testing. ^&^18 microbes were fully concordant, though one was defined as concordant despite not being adjudicated as a cause of pneumonia when BALF-mcfDNA data were available to adjudicators. For 7 concordant detections, BALF-mcfDNA sequencing identified the species where composite-UC testing identified only the genus of the causative microbe; *for illustration, one UC detection was counted twice because UC detected *Aspergillus sp.,* whereas two species of *Aspergillus* were detected by BALF-mcfDNA. 10 adjudicated taxa were detected only by composite-UC testing. All microbe details in Tables S9-12.

#### Clinical specificity

Of 118 samples, 38 (32%) had no reported microbes, 39 (33%) had 1-3, and 41 (35%) had ≥4 (mean 2.7 per report; Fig. 5A), some representing likely respiratory commensals (16). To aid with clinical interpretation, the mcfDNA sequencing test classifies all reported taxa as *Obligate and Opportunistic Pathogens*, *Microbes with Pathogenic Potential and DNA viruses*, or *Upper Respiratory Tract Flora*. Overall, 8.5% of detections were *Obligate and Opportunistic Pathogens*, 27.6% were *Microbes with Pathogenic Potential and DNA viruses*, and 63.9% were *Upper Respiratory Tract Flora* (Fig. 5B). Adjudication determined a lung infection etiology for 79% of *Obligate and Opportunistic Pathogen* detections, compared with 28% and 11% for the other two groups (Fig. 5C).

**Figure 5.**
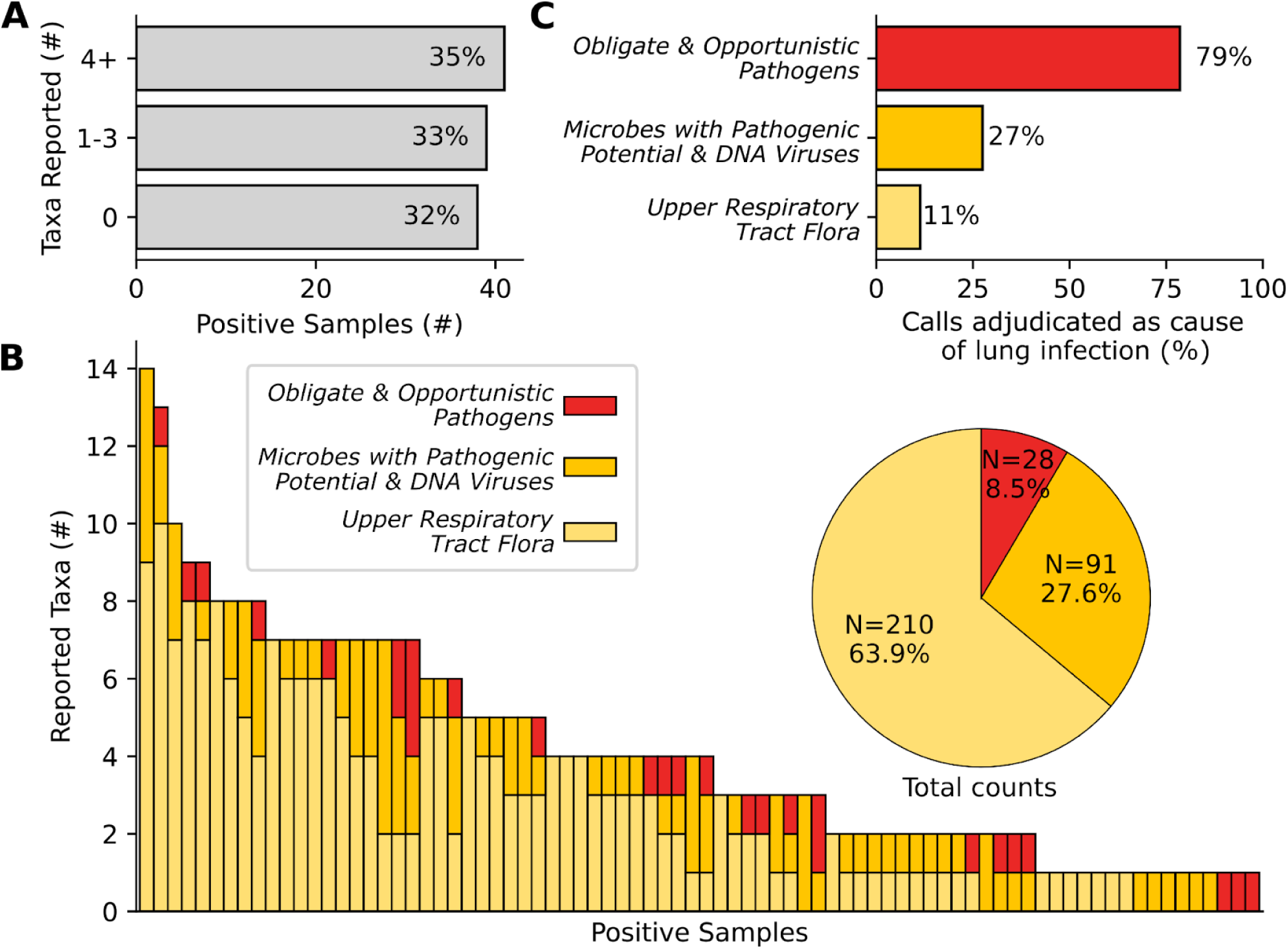
Specificity of BALF-mcfDNA sequencing. **A**. The number of microbes per BALF-mcfDNA test report. **B**. Most reported taxa are common commensal flora of the upper respiratory tract. Each vertical bar represents a patient sample. The pie chart shows the total counts of reportable taxa per category. **C**. Microbe categories correlate with different likelihood of causing pneumonia.

Clinical specificity was assessed in the 73 samples with no adjudicated cause of pneumonia. At least one taxon was reported in 37 of 73, largely *Upper Respiratory Tract Flora* (100/121 detections) or *Microbes with Pathogenic Potential and DNA viruses* (19/121); only two detections were *Obligate and Opportunistic Pathogens*. Per-taxon specificity exceeded 95% for 511/522 CRR tax, with 489 showing 100% specificity (Fig. S6). The 11 taxa with <95% specificity included ten Upper Respiratory Tract Flora and one Microbe with Pathogenic Potential & DNA Virus. BALF-mcfDNA quantitation BALF-mcfDNA sequencing quantifies the mcfDNA concentration from each microbe. Microbes adjudicated as the cause of pneumonia had significantly higher concentrations than non-causative microbes (Fig. 6A). Positive predictive value (PPV; percent of causative microbes) increased with higher concentration thresholds across all categories (Fig. 6B), indicating that higher concentrations correlate with pneumonia causality in the clinical adjudicated gold standard. PPV estimates at very high thresholds were unreliable with few high-concentration detections and some above ULoQ.

**Figure 6.**
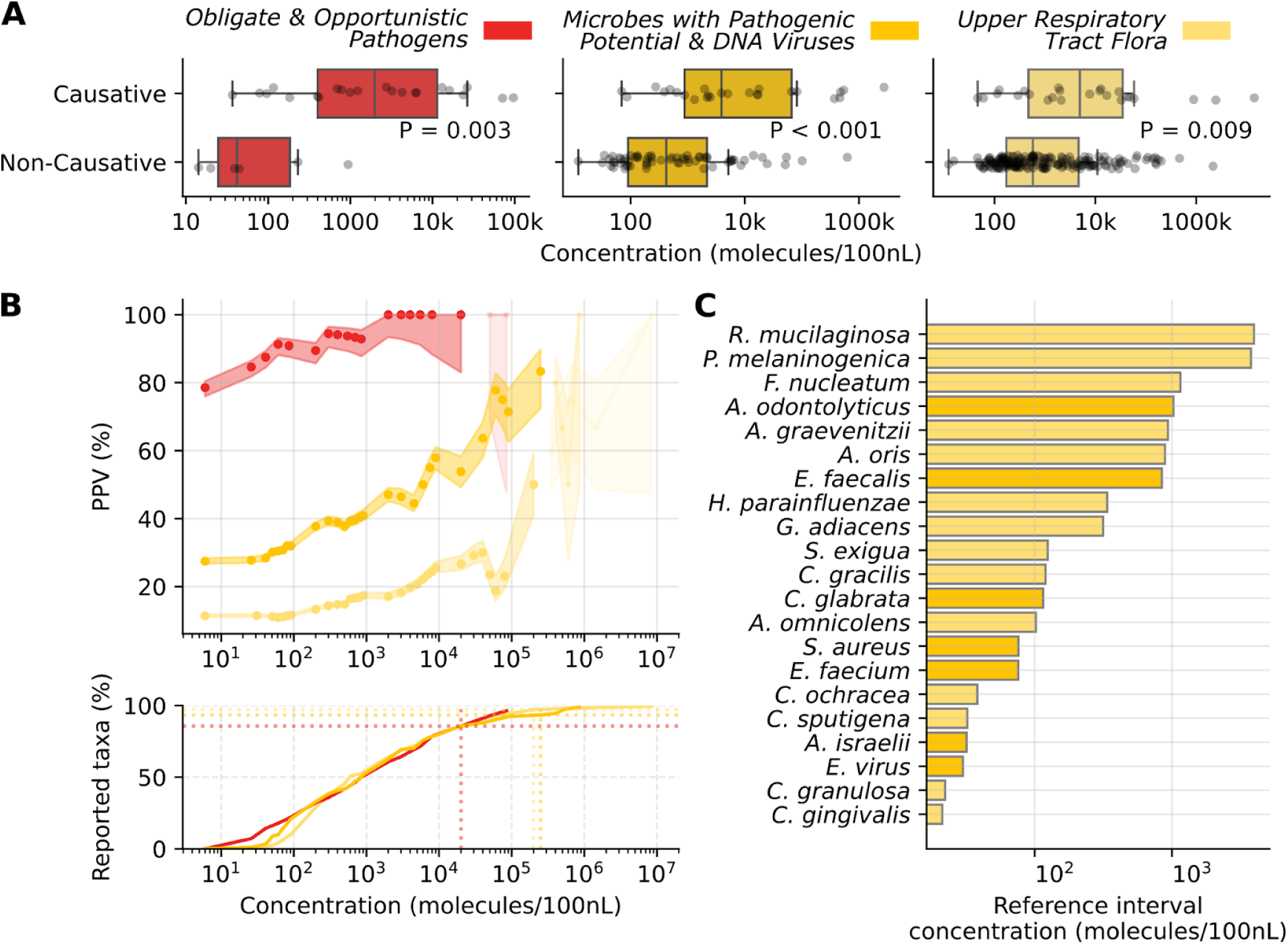
Clinical relevance of microbial quantification. **A**. Microbes adjudicated as the cause of pneumonia (Causative) are detected at a significantly higher concentration than other microbes (Non-Causative) (P < 0.05; Mann-Whitney test for each category). Black points represent individual detections. **B**. (upper panel) The positive predictive value (PPV) was assessed as the fraction of all reported microbes that are causative at a range of concentration thresholds; for each threshold, detections at or above the threshold are considered. The 95% confidence intervals at each threshold are shown as the shaded area around the line. Low-confidence data are defined as thresholds where the difference between the lower and upper 95% CIs exceeds 20% (lighter shading; higher concentration thresholds with fewer reported taxa). (lower panel) Most reported taxa coincide with the high-confidence range of concentration values. The cumulative percent of reported taxa below the maximum high-confidence thresholds are 96%, 93% and 97% of *Obligate & Opportunistic Pathogens*, *Microbes with Pathogenic Potential & DNA Viruses* and *Upper Respiratory Tract Flora* taxa, respectively. **C**. Reference intervals were calculated for 15 taxa with sufficient non-causative detections. 10/15 reference interval taxa were *Upper Respiratory Tract Flora*.

Reference intervals, based on the 97.5th percentile of non-causative microbe concentrations (healthy BALF unavailable), were set for all CRR microbes: 501/522 taxa had a reference interval of 0 molecules/100nL, as they were reported in ≤2.5% of specimens, and 21/522 taxa had non-zero intervals (Fig. 6C).

## Discussion

The comprehensive analytical validation of the BALF-mcfDNA test has addressed the many challenges in validating a test with such a wide range of microbes, through the design and use of a panel of representative microbes that span the range of genome size and GC content of taxa with potential to cause lung infections. Experiments using these contrived samples were performed alongside experiments in clinical samples, to demonstrate properties of the test.

Using *in-silico* experiments, we probed the impact of natural variation and co-infection on reported results, addressing details which could not be obtained using contrived or clinical samples. The validation experiments demonstrated that BALF-mcfDNA sequencing is sensitive, specific, robust to natural genetic variation, and provides both accurate and precise quantitative concentration estimates.

Clinical validation showed that BALF-mcfDNA sequencing is a compelling supplement to UC for diagnosing lung infections. BALF-mcfDNA sequencing demonstrated a diagnostic yield that was 43% higher than UC testing, and when combined with UC testing, diagnostic yield increased by 57%. This demonstrates the strong complementary value of this diagnostic approach for immunocompromised pneumonia, which is a highly morbid, often fatal condition with limited effective diagnostics despite extensive existing tests. BALF-mcfDNA sequencing uniquely identified pathogens such as *Nocardia spp., Aspergillus spp., Mucorales, Legionella spp.,* and anaerobes, organisms often missed by UC and not covered by standard antimicrobial therapy. Failure to diagnose and treat such organisms can result in sub-optimal clinical outcomes, including death. In over half of cases with concordant UC and mcfDNA findings, the test either identified additional likely causes of infection or provided species-level fungal identification, not achieved by UC. These findings align with recent work showing that mcfDNA-based metagenomic sequencing of BALF outperforms both UC and whole-cell DNA methods for diagnosing pulmonary infections, supporting its use as a first-line diagnostic tool(17). Unlike earlier proof-of-concept studies(18–20), the BALF-mcfDNA test reported here (Karius Focus|BAL) is commercially available, delivering results within 24 hours of sample receipt.

Although BAL introduces sterile saline, BALF may contain DNA from upper airway flora that are more often commensals than pulmonary pathogens. In culture-based testing, such organisms are typically grouped and reported as normal respiratory flora. To improve specificity, and help interpretability, the reportable taxa for Karius Focus|BAL are limited to >500 clinically curated pulmonary pathogens, and the report categorizes taxa by their likelihood of causing lung infection. Interpretation is further supported by quantitative measurements of mcfDNA, accurate across several orders of magnitude, which may distinguish low-level colonization from clinically meaningful microbial presence. The report also presents obligate and opportunistic pathogens as a distinct category, enhancing clinical interpretability. Together, this framework should aid clinicians in contextualizing and interpreting complex microbial detections in BALF.

Plasma-mcfDNA sequencing previously identified a pneumonia etiology in 28% of immunocompromised patients(2). In a subset of the same cohort, BALF-mcfDNA sequencing identified an etiology in 34% of participants. Importantly, plasma mcfDNA testing can also capture infections beyond the lung(2) and may be particularly useful when bronchoscopy is delayed or contraindicated, when concurrent non-pulmonary infection (common in immunocompromised patients(21)) is suspected, or when syndrome-driven testing is less reliable. These results underscore the complementary roles of plasma- and BALF-based mcfDNA sequencing in evaluating suspected lung infections in immunocompromised patients.

This study has several limitations. While P11 taxa in contrived samples represent the CRR’s microbial scope, performance may vary for untested taxa. Clinical and study samples were stored rather than freshly collected, which may impact performance. Inference between very closely related taxa may yield ambiguous detections, as shown *in-silico*. Some low-burden pathogens (e.g. *Aspergillus*) are difficult to diagnose by any modality, indicated by missed detections in both UC and mcfDNA sequencing (Tables S9 and S10), suggesting the complementary nature of different testing methods. Characterization of upper respiratory flora in BALF is complex, with adjudicators disagreeing about the interpretation of some results (Fig S6 and Table S9). Quantitative data for each taxon were available to adjudicators; though not formally used in assessments, they may have influenced adjudicator judgments. Finally, prior PICKUP adjudications were retained even when alternative causes were identified by BALF-mcfDNA, potentially overestimating UC-testing performance.

In conclusion, this study demonstrates that BALF-mcfDNA sequencing represents a significant advancement in the diagnosis of lung infections, offering sensitive, comprehensive pathogen detection, and timely results compared to conventional testing methods. These data suggest that BALF-mcfDNA sequencing could become an integral part of the standard diagnostic workflow for pulmonary infections, potentially leading to improved patient management.

## Ethics approval

All participants in the PICKUP study provided written informed consent. Ethical review and oversight were conducted by Advarra. The ethical committees of all participating institutions gave ethical approval for this work.

## Data Availability

All data produced in the present study are available upon reasonable request to the authors.

## Acknowledgements

We thank the following Karius teams; Laboratory Operations for testing the plasma and BALF samples, Clinical Affairs for clinical data management and analysis, Molecular Biology for assay development, Medical Affairs for adjudicator training, and the Scientific Review committee for their thoughtful review of the manuscript. We thank Dr. LauraLe Dyner, Medical Affairs, for contributing to the clinical curation of the BALF CRR. We also thank the independent panel for adjudicating the probable causes of pneumonia: Drs. Bert Lopansri, University of Utah; Christopher Cooper, Advent Health Orlando; Madeleine Heldman, Duke University School of Medicine; Megan Morales and Nicole Vissichelli, Virginia Commonwealth University School of Medicine; Rosy Kodiyanplakkal, Weill Cornell Medicine; Sanjeet Dadwal, City of Hope National Medical Center; Uriel Sandovsky, UT Southwestern Medical Center; Kiran Gajurel, Wake Forest University School of Medicine; Michael Angarone, Northwestern University and Sharjeel Ahmad, University of Illinois College of Medicine-Peoria. During the preparation of this work the authors used ChatGPT in order to condense text and reduce the word count of some sections. After using this tool/service, the authors reviewed and edited the content as needed and take full responsibility for the content of the published article.

This article has an online data supplement, which is accessible from this issue’s table of content online at www.atsjournals.org

## Author contributions

Kevin Brick : Accessed and verified data, Data curation, Formal analysis, Investigation, Methodology, Validation, Visualisation, Writing original draft, Writing review & editing

Kristin H. Jarman : Accessed and verified data, Conceptualisation, Data curation, Formal analysis, Investigation, Methodology, Validation, Visualisation, Writing original draft, Writing review & editing

Frederick Nolte : Conceptualisation, Investigation, Methodology, Writing original draft, Writing review & editing

Igor D. Vilfan : Formal analysis, Investigation, Methodology, Project administration, Resources, Supervision

Marla Lay Vaughan : Investigation, Methodology, Resources, Validation

Aga Zielinska : Conceptualisation, Funding acquisition, Project administration, Resources, Writing original draft

Victoria Portnoy : Investigation, Methodology, Validation

Bradley A. Perkins : Conceptualisation, Funding acquisition, Supervision, Writing review & editing

Sivan Bercovici : Conceptualisation, Funding acquisition, Methodology, Supervision, Validation, Writing review & editing

Timothy A. Blauwkamp : Conceptualisation, Funding acquisition, Methodology, Resources, Supervision, Validation, Visualisation, Writing original draft, Writing review & editing

Jason D. Goldman : Resources, Formal analysis, Supervision, Validation, Writing review & editing

Joshua A. Hill : Resources, Formal analysis, Supervision, Validation, Writing review & editing

Stephen P. Bergin : Accessed and verified data, Conceptualisation, Resources, Supervision, Writing review & editing

