## Supplementary information for "Microbial Cell-Free DNA Sequencing for Diagnosing Lung Infections in Immunocompromised Patients"

**Running title:** Microbial cfDNA Sequencing in Lung Infection

**Subject category:** Diagnosis of Infections

**Supplementary methods** **4**
Bronchioalveolar lavage microbial cell free DNA (Karius FocusTM|BAL) 4
Sample preparation and sequencing 4
Clinical Reportable Range (CRR) 4
Data processing 4
Analytical validation 5
Reference materials and contrived samples 5
Limit of detection (LoD) and reportable range 5
In-silico simulations 6
Inclusivity, Exclusivity and Informatics Stability 6
Cross-reactivity 6
Accuracy 7
Precision 7
Clinical validation 7
**Supplementary tables** **9**
Table S1. BALF-mcfDNA test Clinical Reportable Range (CRR). 9
Table S2. LoD concentrations for individual and grouped taxa. 9
Table S3. Concentration estimates for all P11 taxa exhibit similar precision. 11
Table S4. BALF-mcfDNA sequencing is robust to natural genetic variation. 12
Table S5. High specificity despite natural genetic variation in off-CRR taxa. 13
Table S6. Taxon pairs exhibiting quantitative interference > 20%. 14
Table S7. Patient Characteristics 17
Table S8. Case data for clinical validity study. 18
Table S9. Microbes determined as the cause of pneumonia in clinical patients in which the only adjudicated causes were determined from the BALF-mcfDNA test. 19
Table S10. Microbes determined as the cause of pneumonia in clinical patients in which only composite-UC testing identified an adjudicated cause. 20
Table S11. Microbes determined as the cause of pneumonia in clinical patients in which BALF-mcfDNA sequencing identified additional adjudicated taxa. 21
Table S12. Microbes determined as the cause of pneumonia in clinical patients in which both composite-UC testing and BALF-mcfDNA sequencing identified the same adjudicated cause(s) and in which BALF-mcfDNA sequencing did not identify additional adjudicated causes. 22
**Supplementary figures** **24**
Figure S1. LoD is not correlated with genome GC content or length. 24
Figure S2. Impact of high human concentrations on LoD. 25
Figure S3. Linear quantification for individual taxa. 26
Figure S4. Variation in DNA concentration estimates of P11 taxa using automated electrophoresis. 28
Figure S5. Criteria for exclusion of enrolled BAL specimens from the clinical validation. 29
Figure S6. Most CRR taxa have high specificity. 30
**Supplementary references** **31**

### Supplementary methods

**Bronchioalveolar lavage fluid microbial cell free DNA sequencing (Karius Focus™|BAL)**

***Sample preparation and sequencing***

Thawed BALF and control samples were spiked with internal controls (synthetic ID sequences ("ID spikes") to monitor contamination and whole-assay internal normalization control (WINC) molecules to assess efficiency(1)). Libraries were prepared using the Helion in-matrix method(2) on Hamilton STAR liquid handlers, pooled with four no-template environmental controls (EC) and two assay controls (AC; positive controls containing DNA from *Aspergillus fumigatus, Escherichia coli,* and *Pseudomonas aeruginosa*), and sequenced on Illumina NextSeq500/550 or NovaSeq6000 (~25 million reads/sample). QC required ≥1,000,000 estimated deduplicated reads (EDR), acceptable control performance, and absence of contamination as detected via ID spikes(1). Batch QC further required correct EC/AC results, with reprocessing or review for any failures. McfDNA concentration was reported as molecules/100nL per microbe.

***Clinical Reportable Range (CRR)***

From a curated subset of >1,000 taxa (Karius Spectrum™ test CRR(1, 3)), a board-certified infectious disease physician (LLD) and clinical microbiologist (FSN) retained >500 taxa potentially causing lung infections, based on reference texts(4, 5) and literature review. This set is deemed the Clinical Reportable Range (CRR; Table S1). Reported microbes were functionally categorized as:

- Obligate and Opportunistic Pathogens: May cause lung infection.

- Microbes with Pathogenic Potential and DNA Viruses: May cause lung infection or represent commensals.

- Upper Respiratory Tract Flora: More commonly commensals than the cause of lung infection.

***Data processing***

Metagenomic sequencing data was processed similar to previously described(1). Reads were demultiplexed (bcl2fastq v2.20.0.422-2(6)), trimmed (Trimmomatic v0.32(7); >20 bp retained), and aligned to human (GCF_000001405.38), pig (GCA_000003025.6), decoys (GCA_000212995.1, GCA_000786075.2 & GCA_000442295.1) and synthetic references (Bowtie2 v2.2.4(8)) for removal. Residual reads underwent k-mer filtering to exclude human satellite DNA, then aligned to a curated microbial database(1) (BLAST v2.15.0(9)). After filtering mitochondrial, plasmid, and control-derived reads, PCR duplicates were removed. Taxon abundance was estimated from alignments and refined by expectation-maximization to yield estimated deduplicated reads (EDR)(1). mcfDNA concentration was derived from EDR and WINC counts(1). Batch EC samples modelled environmental EDRs and statistical significance was computed per taxon based on EC EDRs, with separate thresholds for low and regular EC-risk taxa (EC risk assessed from commercial clinical plasma-mcfDNA sequencing data). Significant CRR taxa were checked for uniform coverage and cross-reactivity. Taxa passing all filters were reported.

#### Analytical validation

##### *Reference materials and contrived samples*

Eleven CRR microorganisms were chosen that spanned the range of genome size and GC content of CRR taxa. Equimolar amounts of genomic DNA from these microorganisms (ATCC reference genomes), fragmented to reflect mcfDNA fragment lengths in BALF (NEB Ultra-II FS, New England Biolabs), were combined (P11-mixture). A commercially acquired BALF matrix (University of Pittsburgh Medical Center biorepository) with no reported P11 microbes (based on pre-validation BALF-mcfDNA) was spiked with P11-mixture. Contrived LoD samples used a matrix diluted to $3.3 \times 10^6$ human mcfDNA molecules/100nL (10th percentile of pre-validation BALF), and Precision samples used $1.1 \times 10^7$ molecules/100nL (median).

##### *Limit of detection (LoD) and reportable range*

Following CLSI EP17-A2, P11 was spiked into BALF at 13 concentrations (0.1 to 20,000 molecules/100nL), with 20 replicates per dilution, from four reagent lots. Running batches on separate days, the LoD was calculated using probit analysis for each P11 taxon at five sequencing depths (10%, 25%, 50%, 75%, 90%) expected in clinical samples. The weighted average across down-sampled sequencing depths was calculated (global LoD). The reportable range was defined by the lower and upper limits of quantitation (LLoQ/ULoQ)(1). LLoQ was the lowest concentration with a coefficient of variation (CV) <50%. ULoQ was the maximum concentration in the range of concentrations tested where the correlation coefficient of spiked vs. measured concentration is >0.95.

##### *In-silico simulations*

*In-silico* samples were used to assess robustness, inclusivity, exclusivity, and cross reactivity. Reads were simulated from chosen assemblies with lengths sampled from pre-validation clinical BALF samples. Simulated reads were added to a FASTQ of actual sequencing reads from a BALF sample, and downsampled to median depth. For inclusivity and exclusivity, the pipeline was blinded to simulated assemblies to assess read alignment for taxa with natural genetic diversity. Genomic distances were computed between assemblies by performing pairwise comparisons using a modified MinHash algorithm(10).

##### *Inclusivity, Exclusivity and Informatics Stability*

To assess inclusivity, *in-silico* samples were generated for 20% of CRR taxa (10% taxa from historical samples; 10% random taxa; N=106). Only taxa with multiple assemblies were tested. A randomly selected assembly was spiked at twice global LoD and the genomic reference database was blinded to the spiked assembly to model reads from a strain not present in our database. Inclusivity is the percent of simulated taxa that were reported.

To assess exclusivity, *in-silico* samples were generated for selected microbes not included in the CRR (50% shared a parental taxon with on-CRR taxa; 50% randomly selected from clinical samples; N = 106). Taxa with >1 assembly were spiked at median concentrations (pre-validation samples), and the genomic reference database was blinded to the spiked assembly to model interference from a strain not included in the CRR.  Exclusivity is the percent of samples where no taxa were reported.

To assess informatics stability, *in-silico* samples were generated (5,000 reads) for each CRR taxon. No database blinding was performed, as the goal was to assess if the full pipeline can detect all CRR taxa.

##### *Cross-reactivity*

*In-silico* samples were generated with simulated reads from two related taxa. We selected 80 pairs of assemblies (α1, α2), chosen from taxa with >1 assembly and commonly observed in plasma-mcfDNA clinical samples. For each pair, seven samples were generated using different counts of simulated reads for α1 and α2, respectively, as follows: [5000;0], [15000;0], [0;5000], [0;15000], [5000;5000], [5000;15000], [15000;5000]. Cross-reactivity was defined as the difference between expected (assembly spiked alone) and observed EDRs.

##### ***Accuracy***

Quantitative accuracy compared automated electrophoresis (TapeStation, Agilent, Santa Clara, CA) DNA concentration estimates to estimates from BALF-mcfDNA sequencing. Replicate dilution-series samples from the LoD experiment were used, which satisfy CLSI EP09-A3(11). Variance in automated electrophoresis estimates was calculated from 8-10 independent P11 library measurements across different days (Fig. S4). For each taxon, the percentage difference from the median was calculated, and a randomly selected difference was applied to each electrophoresis estimate to simulate expected variation. Quantitative accuracy was the average difference between automated electrophoresis and BALF-mcfDNA concentration estimates.

##### ***Precision***

Precision assessment followed CLSI EP05-A3 guidelines(11). Within-run and between-run precision were assessed using P11 spiked into human BALF reference matrix at concentrations above LoD (60x & 200x global LoD). Sequencing batches used three reagent lots run by 8 operators on two instruments. Repeatability was the median CV of within-day triplicates; reproducibility was calculated between days. Clinical precision was assessed on taxa reported at a concentration above LoD in BALF reference matrix samples that were run in triplicate on eight separate days; pairwise call concordance measured qualitative precision.

#### Clinical validation

Clinical validity was evaluated through retrospective analysis of data from a prospective, multicenter study at 10 U.S. tertiary care centers (PICKUP study, NCT04047719)(12, 13). Hospitalized patients undergoing diagnostic bronchoscopy for suspected pneumonia were eligible if they were receiving treatment for active hematologic malignancy, recently underwent hematopoietic cell transplantation, or were receiving immunosuppressive therapy for treatment of graft versus host disease. Eligible patients underwent bronchoscopy ≤1 day prior or were scheduled for bronchoscopy within 5 days of enrollment. Patients with pre-established pneumonia etiology, recent COVID-19, or insufficient BALF volume were excluded. Of 249 eligible participants from the original study(12), 118 met our inclusion criteria (Fig. S5).

Participants received the protocol-required minimum diagnostic standard plus additional site-specific testing during routine care to identify pneumonia pathogens. The minimum diagnostic standard included bacterial/fungal/mycobacterial cultures and *Pneumocystis jirovecii* stain or molecular testing from BALF, blood culture, serum galactomannan, and respiratory viral panel testing from nasal swab or BALF. The composite of the minimum diagnostic standard and all additional site-specific testing is henceforth deemed usual care (UC) testing. BALF-mcfDNA sequencing results were compared to UC results from BALF to determine diagnostic sensitivity, specificity and additive diagnostic value. Nine independent adjudicators with experience diagnosing and managing immunocompromised patients with lung infections identified the probable microbial cause of pneumonia. Adjudicators were provided with UC results, BALF-mcfDNA results, plasma-mcfDNA results, medical history and concurrent conditions, relevant clinical exam findings, consultation notes, imaging reports, and details of anti-infective medications. The causes of pneumonia in all subjects were previously adjudicated from UC and plasma mcfDNA test results, but without BALF-mcfDNA results(12). Cases where BALF-mcfDNA did not identify new microbes were not re-adjudicated. All original adjudications were retained, even when BALF-mcfDNA sequencing identified a more likely cause of pneumonia. Adjudications based solely on non-BALF UC testing were excluded. In the re-adjudication for this study, cases were independently reviewed by two adjudicators; disagreements required a third adjudicator. Adjudicators were asked to determine if each BALF-mcfDNA detection was a probable cause of pneumonia and to detail what supported each adjudication.

### Supplementary tables

Note: Table S1 is provided as a separate file

**Table S1. BALF-mcfDNA test Clinical Reportable Range (CRR).**

| | Sequencing depth (percentile in clinical samples) | | | | | |
|---|---|---|---|---|---|---|
| | **10%** | **25%** | **50%** | **75%** | **90%** | **Global** |
| **A. fumigatus** | 348 | 239 | 134 | 78 | 30 | 163 |
| **MTB complex** | 236 | 116 | 132 | 86 | 43 | 120 |
| **Low-EC taxa** | 304 | 214 | 133 | 83 | 45 | 153 |
| **B. pertussis** | 883 | 508 | 367 | 205 | 108 | 402 |
| **C. parvum** | 439 | 448 | 378 | 153 | 145 | 323 |
| **E. coli** | 1,571 | 1,108 | 467 | 425 | 420 | 762 |
| **HAdV-B** | 826 | 741 | 394 | 210 | 122 | 462 |
| **L. major** | 1,025 | 439 | 278 | 114 | 152 | 369 |
| **P. aeruginosa** | 925 | 790 | 681 | 463 | 215 | 629 |
| **P. falciparum** | 543 | 400 | 416 | 99 | 139 | 324 |
| **S. aureus** | 420 | 368 | 195 | 119 | 57 | 233 |
| **S. enterica** | 762 | 479 | 215 | 200 | 61 | 331 |
| **Regular taxa** | 1,138 | 771 | 539 | 337 | 248 | 595 |
| | **Data above show LoD concentrations in molecules/100nL** | | | | | |

**Table S2. LoD concentrations for individual and grouped taxa.**

The LoD was determined for each of the 11 taxa in the contrived pathogen mix and for each taxon group (Low EC-risk, Regular) across the range of sequencing depths observed in pre-validation clinical samples. The global LoD was also calculated for each taxon and taxon group (right-most column). Values represent the LoDs, expressed in molecules per 100 nanoliters of BAL fluid.

| | Concentration at 60x LoD (molecules/100nL) | | Concentration at 200x LoD (molecules/100nL) | |
|---|---|---|---|---|
| | **repeatability** | **reproducibility** | **repeatability** | **reproducibility** |
| **Aspergillus fumigatus** | 5.0% | 11.7% | 8.7% | 13.1% |
| **Mycobacterium tuberculosis complex** | 6.0% | 14.2% | 9.9% | 15.7% |
| **Bordetella pertussis** | 5.9% | 12.7% | 9.8% | 14.9% |
| **Cryptosporidium parvum** | 6.7% | 13.9% | 8.0% | 13.7% |
| **Escherichia coli** | 4.7% | 12.1% | 8.6% | 12.8% |
| **Human adenovirus B** | 5.1% | 10.1% | 8.0% | 10.2% |
| **Leishmania major** | 7.0% | 17.1% | 9.2% | 17.3% |
| **Plasmodium falciparum** | 8.0% | 17.0% | 8.4% | 15.6% |
| **Pseudomonas aeruginosa** | 5.9% | 13.9% | 9.2% | 15.1% |
| **Salmonella enterica** | 6.1% | 12.9% | 8.7% | 14.0% |
| **Staphylococcus aureus** | 6.9% | 12.7% | 7.7% | 12.1% |

**Table S3. Concentration estimates for all P11 taxa exhibit similar precision.**

(a) Within-run precision (repeatability) and within-laboratory precision (reproducibility) were assessed by measuring mcfDNA abundance in replicate contrived samples (see methods).

| Reported taxa | | N | Details |
|---|---|---|---|
| Expected | Unexpected | | |
| Yes | No | 102 | - |
| Yes | Yes | 1 | *Bordetella parapertussis* reported;<br>Unexpected, related, *Bordetella bronchiseptica* also reported |
| No | No | 1 | *Bacillus cereus* not reported |
| No | Yes | 2 | *Leishmania donovani* not reported;<br>Unexpected, related, *Leishmania infantum* reported |
| | | | *Brucella melitensis* not reported;<br>Unexpected, related, *Brucella abortus* reported |

**Table S4. BALF-mcfDNA sequencing is robust to natural genetic variation.**

The reported taxa in each of the *in-silico* inclusivity simulations are shown. For 102/106 simulations, only the expected taxon was reported. In three simulations, a taxon related to the expected taxon was reported; this can occur when the blinded assembly is more similar to an assembly from a related species than to other assemblies for the species itself. In the simulation in which *Bacillus cereus* was not reported, we found that the simulated reads were predominantly attributed to the closely related *Bacillus bombysepticus*; since this taxon is not on the CRR, it was not reported.

| Status | N | Simulated taxon | Reported taxon |
| --- | --- | --- | --- |
| No reported taxa | 102 | - | |
| One reported taxon | 4 | *Acinetobacter nosocomialis* | *Acinetobacter baumannii* |
| | | *Stenotrophomonas rhizophila* | *Stenotrophomonas maltophilia* |
| | | *Bacteroides finegoldii* | *Bacteroides ovatus* |
| | | *Bacteroides xylanisolvens* | *Bacteroides ovatus* |

**Table S5. High specificity despite natural genetic variation in off-CRR taxa.**

The reported taxa in each of the *in-silico* exclusivity simulations are shown. For 102/106 simulations, no taxa were reported. In four simulations, a taxon related to the expected taxon was reported.

| Simulated taxa | | EDR | | | | |
|---|---|---|---|---|---|---|
| **Target** | **Interferant** | **target spiked** | **interferant spiked** | **exp** | **obs** | **obs/exp** |
| Mycobacterium chimaera | Mycobacterium avium complex (MAC) | 5000 | 15000 | 3268 | 18328 | 5.6 |
| Mycobacterium avium complex (MAC) | Mycobacterium chimaera | 5000 | 15000 | 4769 | 19009 | 4.0 |
| Bordetella bronchiseptica | Bordetella parapertussis | 5000 | 15000 | 4302 | 16752 | 3.9 |
| Aspergillus flavus | Aspergillus oryzae | 5000 | 15000 | 3219 | 11663 | 3.6 |
| Mycobacterium chimaera | Mycobacterium avium complex (MAC) | 5000 | 5000 | 3268 | 8681 | 2.7 |
| Aspergillus oryzae | Aspergillus flavus | 5000 | 15000 | 1884 | 4972 | 2.6 |
| Bordetella bronchiseptica | Bordetella parapertussis | 5000 | 5000 | 4302 | 8731 | 2.0 |
| Mycobacterium avium complex (MAC) | Mycobacterium chimaera | 5000 | 5000 | 4769 | 9558 | 2.0 |
| Aspergillus flavus | Aspergillus oryzae | 5000 | 5000 | 3219 | 6315 | 2.0 |
| Aspergillus oryzae | Aspergillus flavus | 5000 | 5000 | 1884 | 3324 | 1.8 |
| Mycobacterium chimaera | Mycobacterium avium complex (MAC) | 15000 | 5000 | 9711 | 15315 | 1.6 |
| Aspergillus flavus | Aspergillus oryzae | 15000 | 5000 | 9391 | 14355 | 1.5 |
| Bordetella bronchiseptica | Bordetella parapertussis | 15000 | 5000 | 13167 | 17420 | 1.3 |
| Mycobacterium avium complex (MAC) | Mycobacterium chimaera | 15000 | 5000 | 14303 | 18918 | 1.3 |
| Aspergillus oryzae | Aspergillus flavus | 15000 | 5000 | 5785 | 7390 | 1.3 |
| Haemophilus influenzae | Pasteurella multocida | 5000 | 15000 | 2994 | 0 | 0.0 |
| Aspergillus oryzae | Aspergillus terreus | 5000 | 15000 | 2208 | 0 | 0.0 |
| Aspergillus flavus | Aspergillus fumigatus | 5000 | 15000 | 3564 | 0 | 0.0 |

**Table S6. Taxon pairs exhibiting quantitative interference > 20%.**

A zero value for observed EDR indicates that the taxon was not reportable in the sample. The expected EDR (exp) was calculated from a simulated sample spiked only with the target taxon. The observed EDR (obs) was the reported EDR in the interference simulation experiment. Most deviations occur in experiments with one of three highly similar taxon pairs; *Aspergillus flavus/oryzae, Mycobacterium avium/chimerae, Bordetella bronchiseptica/parapertussis.*

| Characteristics | Overall[a] (n = 118) |
|---|---|
| Age, median (IQR), y | 62 (51-69) |
| Female | 36 (30.5) |
| Leukemia | 103 (87.3) |
| Lymphoma | 24 (20.3) |
| Myelodysplastic syndromes | 20 (16.9) |
| Multiple myeloma | 13 (11.0) |
| Transplant | 35 (29.7) |
| Autologous stem cell transplant | 7 (20.0) |
| Allogenic stem cell transplant | 25 (71.4) |
| Chemotherapy w/in 45 d of enrollment | 100 (84.7) |
| Relapse at time of enrollment | 73 (61.9) |
| Remission at time of enrollment | 18 (15.3) |
| Active graft-vs-host disease | 10 (8.5) |
| Immunosuppressive pharmacologic treatment | 9 (7.6) |
| Invasive procedure 1 d before enrollment to day 14[b] | 118 (100) |
| Bronchoscopy[c] | 118 (100) |
| Thoracentesis | 7 (5.9) |
| Transthoracic needle aspiration | 0 (0) |
| Mini BAL | 0 (0) |
| White blood cell count, median (IQR), K/µL | 2.1 (0.5-4.6) |
| Absolute neutrophil count, median (IQR), K/µL | 1.2 (0.1-3.3) |
| Anti-infective medication at enrollment | 118 (100) |
| Antipseudomonal antibacterial | |
| Mold-active antifungal | |
| Anti-MRSA antibacterial | |
| Anti-PJP antimicrobial | |
| Death ≤30 d | 14 (11.9) |
| Overall study mortality | 25 (21.1) |

**Table S7. Patient Characteristics**

Abbreviations: BAL, bronchoalveolar lavage; IQR, interquartile range; MRSA, methicillin-resistant Staphylococcus aureus; PJP, Pneumocystis jiroveci pneumonia. [a]Data are presented as Count (%), or Median (IQR) unless otherwise indicated. [b]Includes invasive diagnostic procedures performed to establish pneumonia etiology. Some patients underwent ≥1 diagnostic procedure. No video-assisted thoracoscopic surgeries were performed in this cohort. [c]One study subject was scheduled for bronchoscopy but did not have the procedure due to declining health status.

Note: Table S8 is provided as a separate file

**Table S8. Case data for clinical validity study.**

| # | Composite-UC adjudication | BALF-mcfDNA adjudication |
|---|---|---|
| ***1*** | - | *Nocardia cyriacigeorgica* |
| ***2*** | - | *Cunninghamella* |
| ***3*** | - | *Rhizomucor pusillus* |
| ***4*** | - | *Pneumocystis jirovecii* |
| ***5*** | - | *Pneumocystis jirovecii* |
| ***6*** | - | *Aspergillus tubingensis* |
| ***7*** | - | *Aspergillus fumigatus* |
| ***8*** | - | *Legionella hackeliae* |
| ***9*** | - | *Legionella pneumophila* |
| ***10*** | - | *Staphylococcus aureus* |
| ***11*** | - | *Pseudomonas aeruginosa* |
| ***12*** | - | *Pseudomonas aeruginosa* |
| ***13*** | - | *Pseudomonas aeruginosa*<br>*Bacillus cereus, Actinomyces oris* |
| ***14*** | - | *Burkholderia cepacia complex* |
| ***15*** | - | *Actinomyces odontolyticus, Haemophilus parainfluenzae, Prevotella melaninogenica, Rothia mucilaginosa* |
| ***16*** | - | *Actinomyces odontolyticus, Fusobacterium nucleatum, Gordonia bronchialis, Granulicatella adiacens, Rothia mucilaginosa, Slackia exigua* |

**Table S9. Microbes determined as the cause of pneumonia in clinical patients in which the only adjudicated causes were determined from the BALF-mcfDNA test.**

Red = *Obligate & Opportunistic Pathogens*; Orange = *Microbes with Pathogenic Potential & DNA Viruses*; Yellow = *Upper Respiratory Tract Flora*.

| # | Composite-UC adjudication | BALF-mcfDNA adjudication |
| --- | --- | --- |
| ***1*** | *Aspergillus species* (only GM+)<br>*Nocardia species* | - |
| ***2*** | *Aspergillus species* (only GM+) | - |
| ***3*** | *Aspergillus species* (only GM+) | - |
| ***4*** | *Mycobacterium species** | - |

**Table S10. Microbes determined as the cause of pneumonia in clinical patients in which only composite-UC testing identified an adjudicated cause.**

Red = *Obligate & Opportunistic Pathogens*; Orange = *Microbes with Pathogenic Potential & DNA Viruses*; Yellow = *Upper Respiratory Tract Flora*. Note that categorizations were not assigned to identifications lacking speciation. GM+ signifies that the sample tested positive in Galactomannan testing. *Mycobacterium species (AFB culture) reported >14 days after enrollment.

| # | Composite-UC adjudication | BALF-mcfDNA adjudication |
|---|---|---|
| ***1*** | *Pneumocystis jirovecii* | *Pneumocystis jirovecii*<br>***Aspergillus fumigatus***<br>***Enterococcus faecium*** |
| ***2**** | *Staphylococcus aureus* | ***Pneumocystis jirovecii*** |
| ***3*** | *Aeromonas caviae* | *Aeromonas caviae*<br>***Citrobacter freundii*** |
| ***4*** | *Aspergillus species* (only GM+)<br>*Staphylococcus aureus* | *Staphylococcus aureus*<br>***Enterococcus faecalis***<br>***Klebsiella pneumoniae*** |
| ***5**** | *Aspergillus species* (only GM+) | ***Actinomyces odontolyticus, Capnocytophaga ochracea, Eikenella corrodens, Enterobacter cloacae complex, Granulicatella adiacens, Prevotella melaninogenica, Pseudomonas aeruginosa, Rothia mucilaginosa*** |
| ***6*** | *Actinomyces graevenitzii* | *Actinomyces graevenitzii*, ***Actinomyces oris, Capnocytophaga ochracea, Fusobacterium nucleatum, Granulicatella adiacens, Prevotella melaninogenica, Rothia mucilaginosa, Slackia exigua*** |

**Table S11. Microbes determined as the cause of pneumonia in clinical patients in which BALF-mcfDNA sequencing identified additional adjudicated taxa.**

Adjudicated taxa identified only by BALF-mcfDNA sequencing are bolded. Red = *Obligate & Opportunistic Pathogens*; Orange = *Microbes with Pathogenic Potential & DNA Viruses*; Yellow = *Upper Respiratory Tract Flora*. Note that categorizations were not assigned to identifications lacking speciation. GM+ signifies that the sample tested positive in Galactomannan testing. *For these samples, the original adjudication (without BALF-mcfDNA sequencing) identified an adjudicated etiology from UC testing, however when BALF-mcfDNA results were provided and the sample readjudicated, taxa identified only by BALF-mcfDNA were adjudicated as likely causes of pneumonia. The taxa adjudicated originally were not detected by BALF-mcfDNA sequencing. Since adjudicators were not asked to reassess UC-based adjudications, it is unknown if BALF-mcfDNA sequencing results would have changed the original adjudications.

| # | Composite-UC adjudication | BALF-mcfDNA adjudication |
|---|---|---|
| **1** | *Aspergillus species* (only GM+) | ***Aspergillus fumigatus*** |
| **2** | *Aspergillus species* (only GM+) | ***Aspergillus fumigatus*** |
| **3** | *Aspergillus species* (only GM+) | ***Aspergillus fumigatus*** |
| **4** | *Mucorales species* | ***Rhizomucor pusillus*** |
| **5** | *Mucorales species* | ***Rhizopus microsporus*** |
| **6** | *Aspergillus species* (only GM+)<br>*Achromobacter xylosoxidans*<br>*Achromobacter denitrificans* | ***Aspergillus niger***<br>***Aspergillus tubingensis***<br>*Achromobacter xylosoxidans*& |
| **7** | *Nocardia abscessus* | *Nocardia abscessus* |
| **8** | *Legionella longbeachae* | *Legionella longbeachae* |
| **9** | *Legionella micdadei* | *Legionella micdadei* |
| **10** | *Cytomegalovirus* | *Cytomegalovirus* |
| **11** | *Staphylococcus aureus* | *Staphylococcus aureus* |
| **12** | *Staphylococcus aureus* | *Staphylococcus aureus* |
| **13** | *Staphylococcus aureus* | *Staphylococcus aureus* |
| **14** | *Pseudomonas aeruginosa* | *Pseudomonas aeruginosa* |
| **15** | *Pseudomonas aeruginosa* | *Pseudomonas aeruginosa* |
| **16** | *Pseudomonas aeruginosa*<br>*Stenotrophomonas maltophilia* | *Pseudomonas aeruginosa*<br>*Stenotrophomonas maltophilia* |
| **17** | *Actinomyces oris*<br>*Stenotrophomonas maltophilia* | *Actinomyces oris* |
| **18** | *Haemophilus parainfluenzae* | *Haemophilus parainfluenzae* |

**Table S12. Microbes determined as the cause of pneumonia in clinical patients in which both composite-UC testing and BALF-mcfDNA sequencing identified the same adjudicated cause(s) and in which BALF-mcfDNA sequencing did not identify additional adjudicated causes.**

In 6 participants (rows 1-6), BALF-mcfDNA sequencing added speciation information to microbes detected by composite-UC (bolded taxa). (&) For the participant in row 6, *Achromobacter xylosoxidans* (detected by both BALF-mcfDNA and UC testing), was not adjudicated as a cause of pneumonia when BALF-mcfDNA sequencing results were seen by adjudicators. Nonetheless, as it was adjudicated as a cause in the UC-only adjudication, it is retained on Figure 4 as a concordant adjudicated detection. Red = *Obligate & Opportunistic*

*Pathogens*; Orange = *Microbes with Pathogenic Potential & DNA Viruses*; Yellow = *Upper Respiratory Tract Flora*. Note that categorizations were not assigned to identifications lacking speciation. GM+ signifies that the sample tested positive in Galactomannan testing.

### Supplementary figures

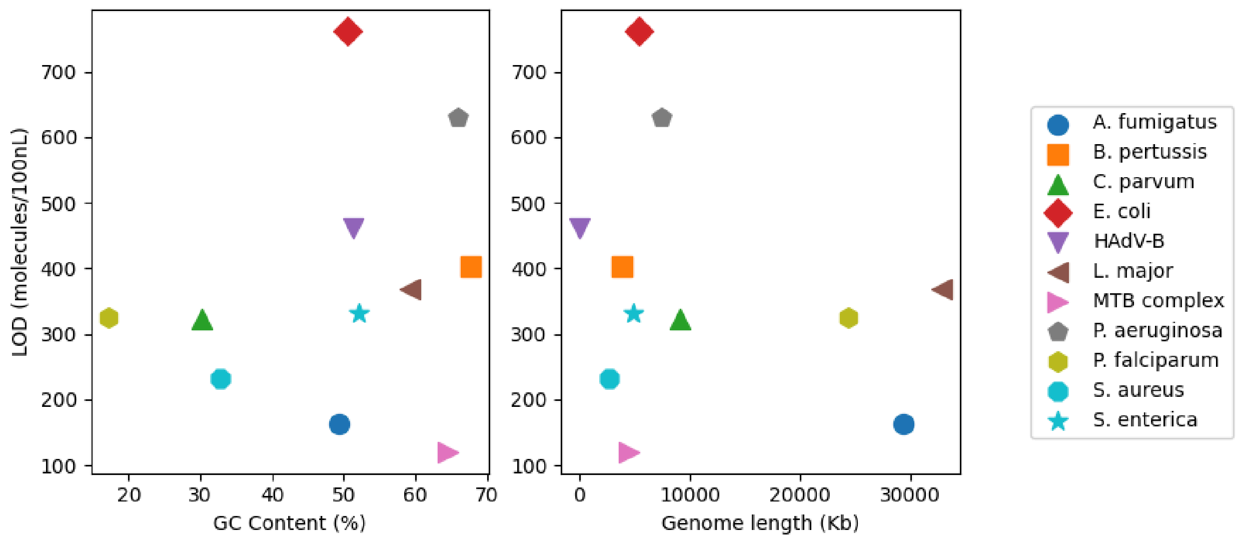

**Figure S1. LoD is not correlated with genome GC content or length.**

The global LoD of each P11 taxon is shown as a function of either genome GC content or genome length. Neither property is a strong correlate of the LoD.

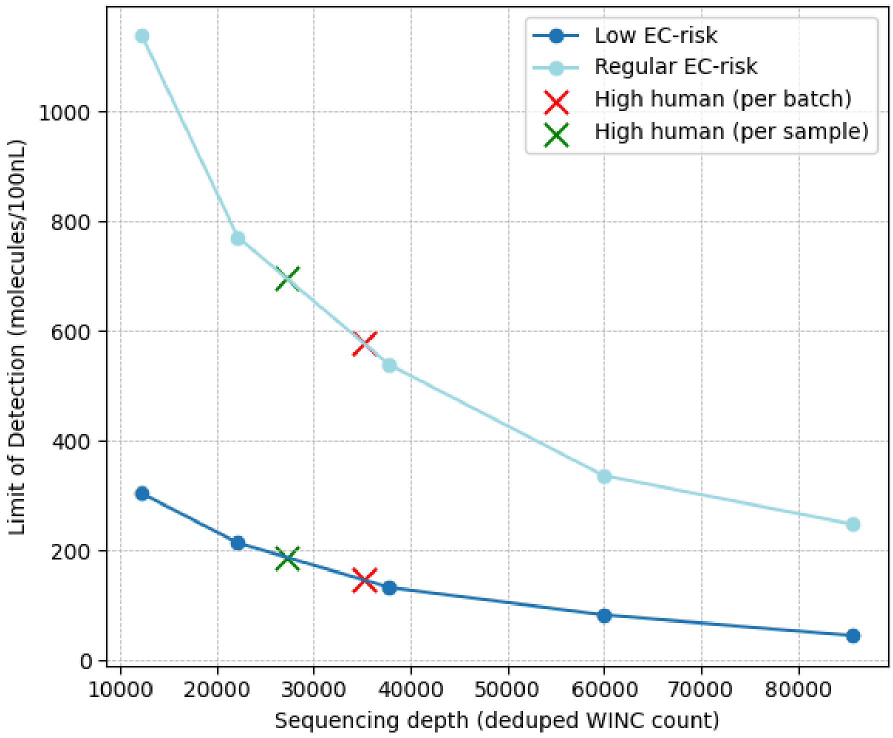

**Figure S2. Impact of high human concentrations on LoD.**

We inferred the LoD for typical sequencing depth that results from the two typical high-human scenarios; i) high per-sample human content (27,172 ddSPANKs; median ddSPANK of samples with high human equivalent to the 90th percentile of human concentrations for samples in clinical validation batches) and ii) high per-batch human content (35,172 ddSPANKs; median ddSPANK of clinical validation batches with 90th percentile batch human concentration). The LoD for each of Low EC-risk and Regular EC-risk taxa were linearly interpolated from the flanking sequencing depths that had been run for the LoD experiment. For high human cfDNA concentration at sample-level, the LoD for regular sensitivity taxa is 697 molecules/100nL (17% above global LoD) and for high sensitivity taxa 188 molecules/100nL (23% above global LoD). For high human cfDNA concentration at batch-level, the LoD for regular sensitivity taxa is 578 molecules/100nL (3% below global LoD) and for high sensitivity taxa 147 molecules/100nL (4% below global LoD).

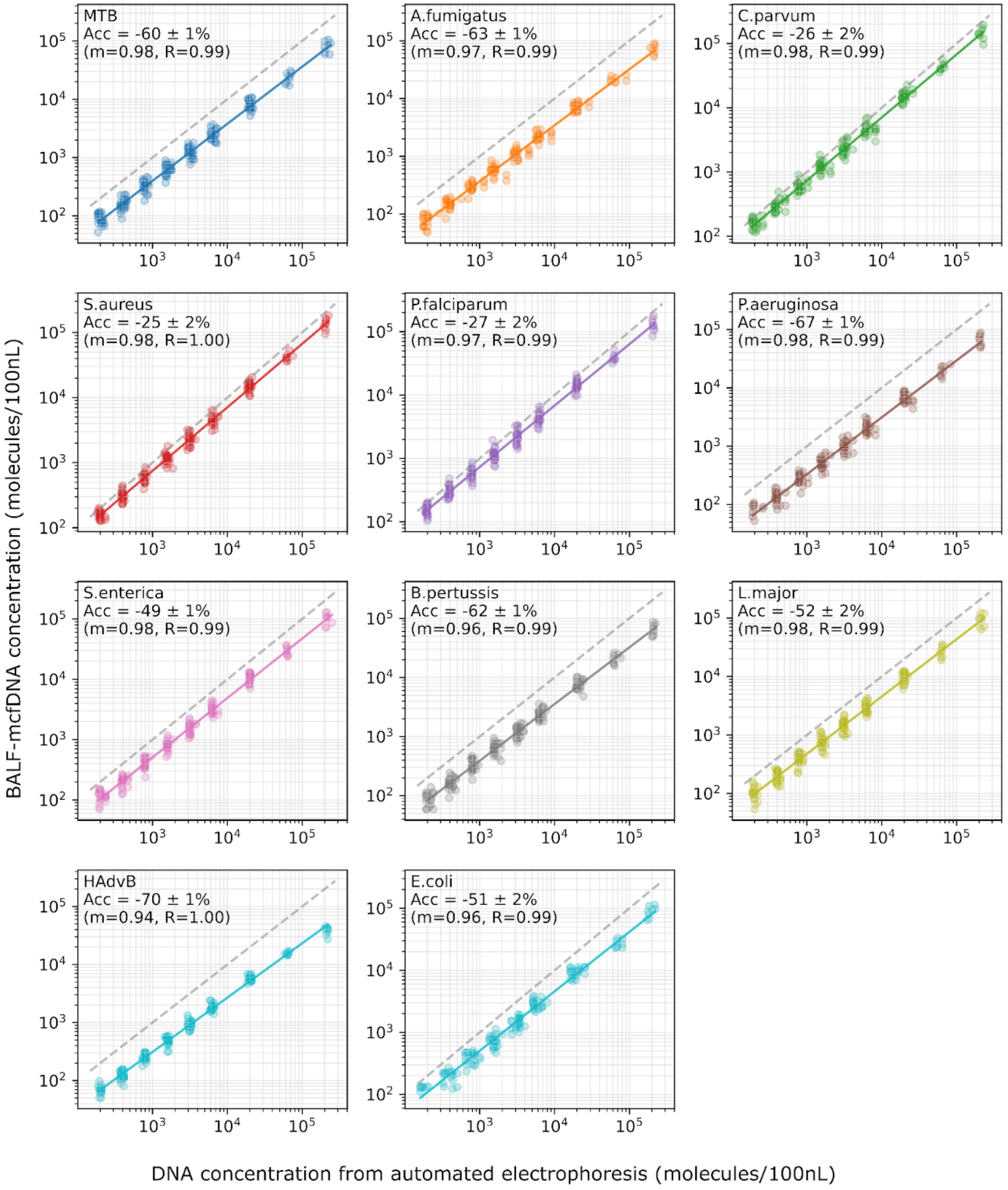

**Figure S3. Linear quantification for individual taxa.**

Individual replicate measurements are shown as circles. The variance observed on the x-axis is from variation in Tapestation measurements that was incorporated in the estimates. The accuracy (Acc), slope of the linear regression line (m) and Pearson correlation coefficient (R) are shown for each taxon. Dashed lines depict a 1:1 regression line with a slope (m) = 1.

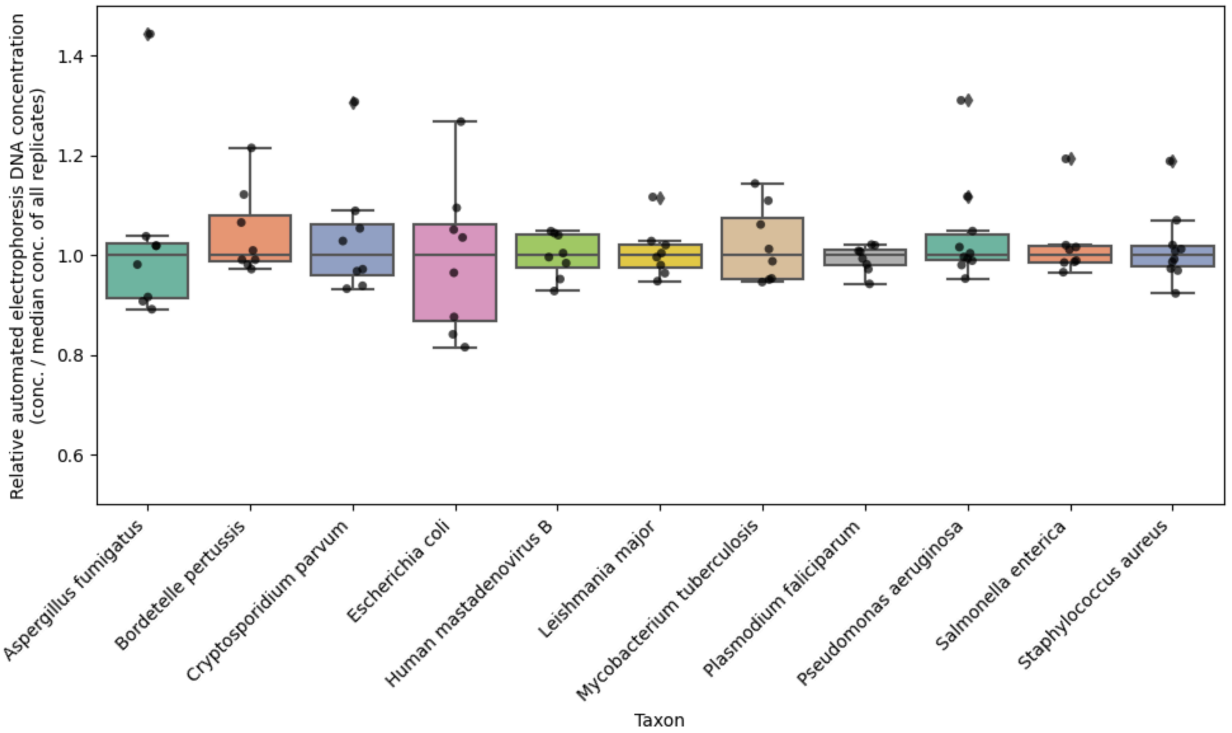

**Figure S4. Variation in DNA concentration estimates of P11 taxa using automated electrophoresis.**

Relative concentration estimates by automated electrophoresis for each of the P11 taxon stocks were taken on five separate days over a 13-day period., taking 2-4 estimates per day. Each point represents one estimate per taxon.

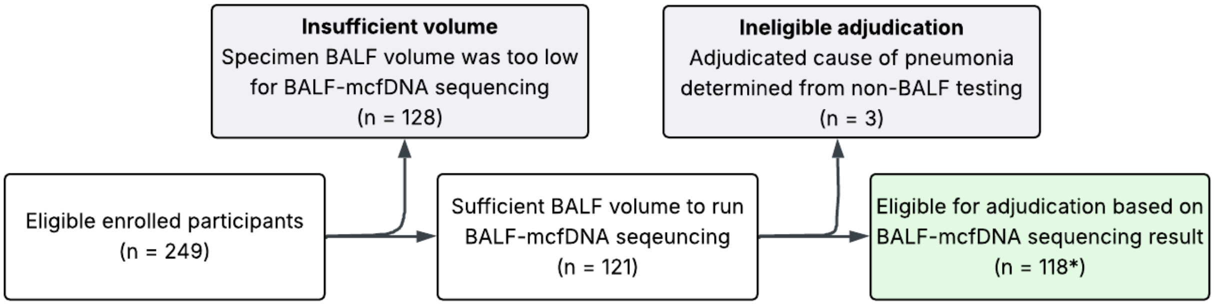

**Figure S5. Criteria for exclusion of enrolled BAL specimens from the clinical validation.**

Specimens with insufficient BALF volume for BALF-mcfDNA sequencing, and specimens with adjudicated causes of pneumonia by usual care testing on non-BALF specimens were removed, resulting in n=118 specimens included in this validation. *Two specimens had potential erroneous adjudications based on a post-study quality review. In both cases, BALF-mcfDNA sequencing detected additional likely causes of pneumonia; [Subject #52] *Aspergillus fumigatus* was detected by BALF-mcfDNA sequencing and also suspected due to acute angle branching of right lung tissue; [Subject #84] *Rothia mucilaginosa* was detected by BALF-mcfDNA sequencing and was detected by bacterial culture and diagnosed by radiography four days prior to enrollment. Both specimens were included in the clinical validity analysis without modification to the official adjudicated causes of pneumonia.

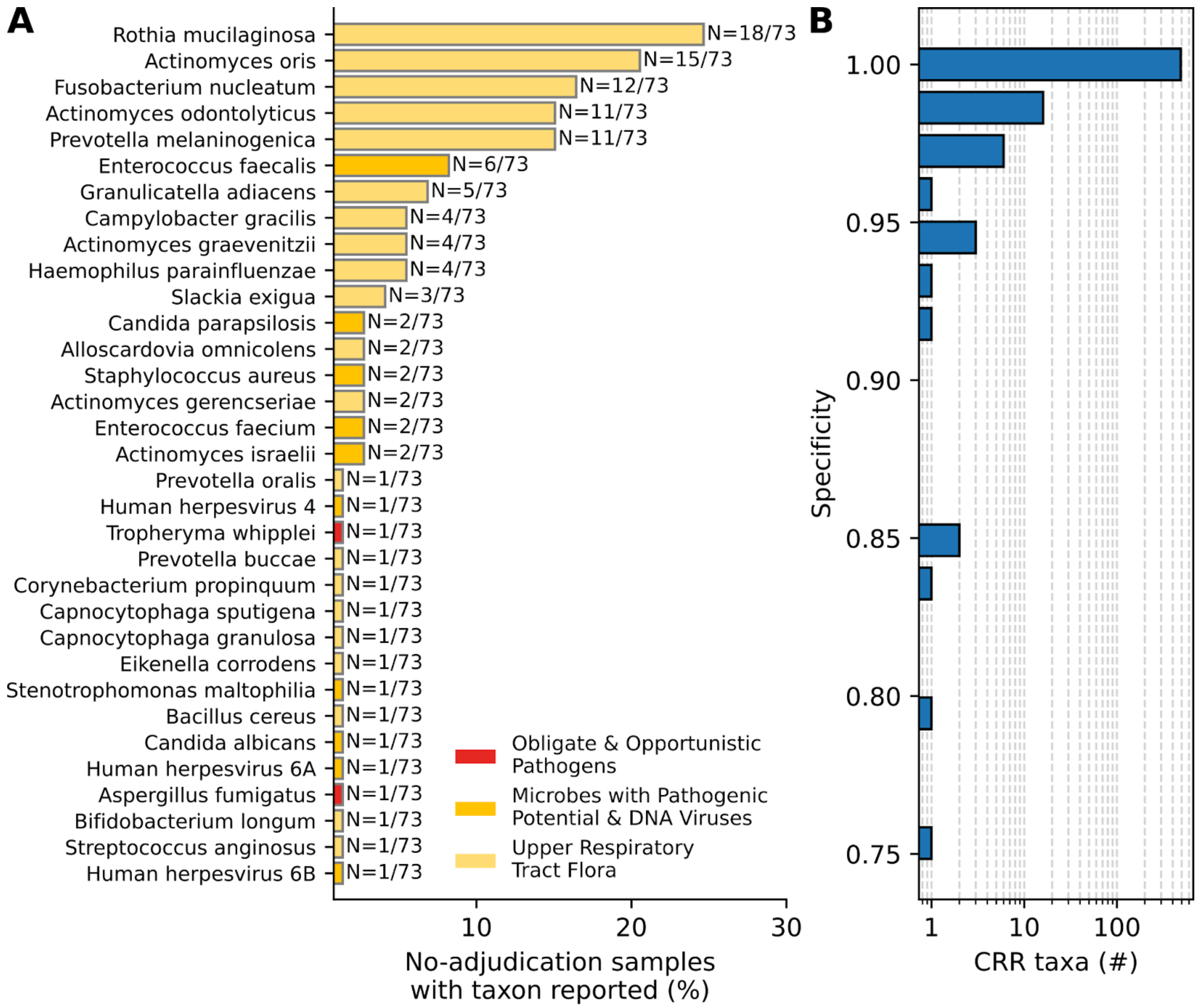

**Figure S6. Most CRR taxa have high specificity.**

Non-adjudicated reported taxa were identified in the 73 samples for which there was no adjudicated cause of pneumonia. These reported taxa were considered False Positive (FP) calls. **A.** Most FP calls were for a small number of predominantly Upper Respiratory Tract Flora. **B.** For each CRR taxon, the specificity was calculated as the fraction of samples without an FP call. Most CRR taxa have 100% specificity.
